# Influence of acute myocardial ischemia on arrhythmogenesis: a computational study

**DOI:** 10.1101/2024.11.20.24317476

**Authors:** Alessandra Corda, Stefano Pagani, Christian Vergara

## Abstract

The early phase of acute myocardial ischemia is associated with an elevated risk of ventricular reentrant arrhythmias. Indeed, after partial or total occlusion of a coronary artery, some regions of the heart experience a reduction in myocardial blood flow. This causes metabolic and cellular processes, such as hypoxia, hyperkalemia and acidosis, which lead to changes in the transmembrane ionic dynamics. The effect of such alterations may result in the formation of electrical loops and reentries.

Computational models could simulate the generation of arrhythmias, possibly persistent, in condition of ectopic beats and in presence of acute myocardial regions. Since quantitative information (extent, localization, …) about acute ischemic regions are hardly available from clinics, to date, computational models only integrate imaging data from chronic infarcted ventricles. This may not accurately reflect the acute condition.

This work presents a novel patient-specific electrophysiological model, based on images of myocardial blood flow maps acquired during a pharmacologically induced acute ischemic event. The model personalization is obtained with the partitioning of the left ventricle geometries on the basis of the myocardial blood flow maps. First, we aim to numerically investigate the induction and sustainment of reentrant drivers in patient-specific scenarios, in order to assess their arrhythmic propensity. Secondly, we perform an intra-patient sensitivity analysis, where different levels of acute ischemia are virtually depicted for the most arrhythmogenic patient. Our results suggest that the amount of ischemic regions seems to have less influence on arrhythmogenesis than their pattern.

## 1 Introduction

Myocardial ischemia is defined as the localized reduction of blood flow and, consequently, oxygen in the cardiac tissue. Most of myocardial ischemia are due to coronary artery disease [1], which is caused by narrowed or completely blocked vessels that are no longer able to provide the physiological blood supply. Depending on how rapidly myocardial ischemia evolves over time, it can be defined in two ways: *acute myocardial ischemia* (AMI), which involves a sudden and significant reduction of blood flow, and *chronic myocardial ischemia*, which develops gradually, often causing persistent symptoms that may worsen over time [2, 3].

Some hours (1.5-5) after the onset of AMI, the histological properties of the myocardium may change resulting in the formation of a scar (myocardial infarct), which can lead to the generation of reentry loops [4]. Other clinically relevant consequences may occur just after the acute ischemic event (first 10 minutes) at the metabolic and ionic level [5, 6]. Indeed, during this phase, *hypoxia* (reduced supply of oxygen) [7], *hyperkalemia* (increased extracellular potassium concentration) [8] and *acidosis* (reduced intracellular pH) [9] occur in the ischemic region. Each of these alterations originates downstream the coronary occlusion and then distributes heterogenously within the myocardium. This subdivision of the myocardium into areas with different ionic and metabolic characteristics causes adverse effects on cardiac electrophysiology, such as heterogeneous action potential morphology and refractory periods: as the ischemic effects worsen, the electrophysiological properties of the tissue will progressively change. Thus, this heterogeneity, induced by acute ischemia, provides an important pro-arrhythmic substrate [10, 11, 6, 12, 13], possibly leading to sudden cardiac death [14]. Specifically, arrhythmias that take place during the first minutes of AMI depend mainly on reentrant mechanisms, contributing to the development of ventricular tachycardia and, possibly, to the degeneration in ventricular fibrillation [10].

The consequences that AMI has on cardiac electrophysiology have been a subject matter of research for many years. Since the processes behind ischemia-induced arrhythmic phenomena in humans are difficulty measurable in the clinical practice, in order to improve their understanding, computational models offer a versatile platform on which virtually explore scenarios that are hardly measurable in the clinic. Pioneering studies performed in 2D geometries allowed to obtain first insights about such processes [15]. First steps toward the computational analysis of the major effects of AMI in 3D geometries was made by studying simplified geometries [16] and animal ventricles with globally ischemic tissue [17], so to characterize the changes in vulnerability to electric shocks during the first phase of AMI. The gradual expansion of the ischemic effects following a coronary occlusion, and consequently the resulting spatial heterogeneity of a human left ventricle, was then introduced in [18, 19, 20]. Because of the difficulty in obtaining AMI data, it is not easy to accurately identify the location and the extent of the ischemic regions. For this reason, the majority of the studies portrayed the heterogeneities of the ionic functions due to AMI through an idealized representation of the ischemic and border zones [21, 22, 23, 24]. More recent studies made the assumption of identifying the acute ischemia regions with infarcted areas reconstructed from imaging, even though it is known that in general they do not match one another [25].

As a consequence, none of the previous studies, to the best of our knowledge, accounted for patient-specific regions characterizing the tissue heterogeneity resulting from AMI. Since the morphology and location of such regions are fundamental to determine arrhythmogenesis in a specific patient, we believe that the use of patient’s AMI regions is fundamental in view of predictive analyses.

The first goal of this work is to overcome the previous limitations, in order to evaluate the arrhythmogenic propensity in human patient-specific acute ischemic scenarios. To this aim, we develop an electrophysiological framework based on a modified version of the Ten-Tusscher Panfilov model [26] that accounts for the ionic variations induced during the early phase of AMI [27]. Specifically, we identify AMI regions, as well as border zones, from patient-specific *perfusion Myocardial Blood Flow* (MBF) maps of four patients suffering from coronary artery disease, where an acute and reversible ischemia has been pharmacologically induced during *stress Computed Tomography Perfusion* (stress-CTP) acquisitions [28]. Arrhythmogenic propensity is numerically studied by means of the 𝕊⊮ *stimulation protocol* applied to different points located in the border region, to simulate patho-physiological conditions where ectopic beats randomly depolarize the tissue.

The second goal of the paper is to assess the influence of the shape and extent of the AMI region on arrhythmogenesis. To this aim, for one specific patient we build virtual scenarios corresponding to different levels of AMI. This allows us to numerically investigate the induction and sustainment of reentrant drivers as a function of realistic acute ischemic scenarios and to assess arrhythmic propensity for different ischemic configurations.

We subdivided the paper into two parts. In the first one we focus on the study of modeling electrophysiology assessing arrhythmogenesis induced by the patient-specific acute ischemia (Methods in Section 2 and Results in Section 3). In the second part we provide a parametric study based on assessing the influence of the dimension and pattern of AMI regions on arrhythmogenesis (Methods in Section 4 and Results in Section 5). Finally, in Section 6 we provide an overall discussion of the outcomes reported in both parts.

## 2 Methods Part I - Arrhythmogenesis in presence of patient-specific acute ischemia

### 2.1 Mathematical model of cardiac electrophysiology

The electrical activation of the heart is the result of two coupled multi-scale processes: at the microscopic scale, the generation of ionic currents through the cellular membrane producing a rapid change of the resting voltage, namely the *action potential*; at the macroscopic scale, the propagation in space of the electrical impulse from cell to cell.

The three-dimensional propagation of the electrical impulse along the cardiac fibers and through the myocardial muscle can be described by means of the *monodomain equation*, a diffusion-reaction partial differential equation in which an assumption of proportionality between the intracellular and extracellular domains is introduced. In this case, the use of the monodomain over more physiologically accurate models, such as the bidomain one [29], is preferred to reduce computational cost since in this work we do not consider external sources such as in defibrillation [30, 31]. This PDE is suitably coupled with a system of ordinary differential equations describing the ionic species concentrations dynamics.

Being Ω the physical domain (i.e., a left ventricle) of the model (See Figure 1, left, where we report as a representative case the geometry of patient M9), at each time *t >* 0 *s* the general form of the coupled problem is as follows: Find **w** ∈ (0, *T*] *×* Ω → ℝ^*d*^ and *u* ∈ (0, *T*] *×* Ω → ℝ, such that:

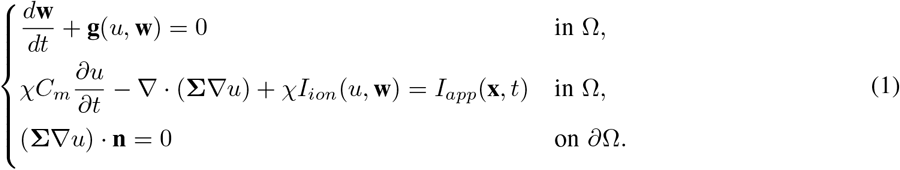

**Figure 1:**
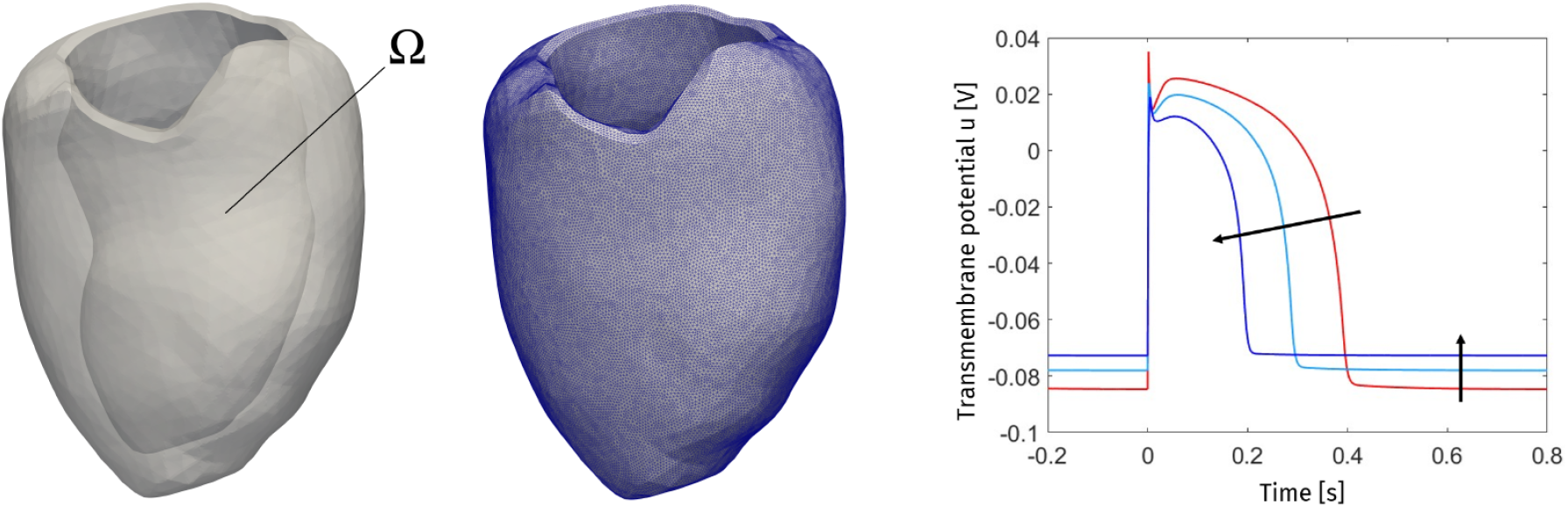
Left: computational domain Ω; Middle: computational mesh; Right: alterations of the action potential for increasing hypoxia, hyperkalemia and acidosis.

The first equation describes the variation over time of the gating variables and of the ionic concentrations **w**, where *u* is the transmembrane potential and **g** is a suitable function depending on the ionic model. The second one is the monodomain equation, where *χ* is the surface area-to-volume ratio, *C*_*m*_ is the membrane capacitance, **Σ** is the anisotropic conductivity tensor, encoding the conduction properties along the the three principal directions. It is in general expressed in terms of three scalar quantities *Σ*_*f*_, *Σ*_*s*_ and *Σ*_*n*_ representing the conductivities along the fiber direction **a**_*f*_, the direction **a**_*s*_ orthogonal to **a**_*f*_ and tangential to sheets, the direction **a**_*n*_ orthogonal to sheets:

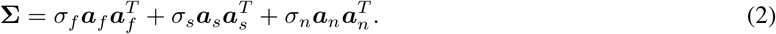

*I*_*ion*_ models the sum of the ionic currents passing through the cell membrane, and *I*_*app*_ accounts for the applied physiological and ectopic impulses. Moreover, homogeneous Neumann conditions are prescribed at the borders of the volume (∂Ω) in order to impose electric insulation. Suitable initial conditions for **w** and *u* should be also prescribed to the system.

### 2.2 Hypoxia, hyperkalemia and acidosis: the modified version of the Ten Tusscher-Panfilov model

When an acute ischemic event occurs, the imbalance between the oxygen demand and supply causes an alteration of the myocardial tissue function: the typical effects are cessation of contraction, alterations in membrane potential, and a variety of metabolic changes. In particular, the major consequences are the shortening of the *Action Potential Duration* (APD) [32, 9] and the increment of the resting potential of the cell (see Figure 1, right), which causes the elongation of the *Effective Refractory Period (ERP)*, i.e. the longest interval between two electrical impulses that fails to depolarize the tissue [33]. At the macroscopic level, *Conduction Velocity* (CV) of the electrical impulse slows down and excitability is reduced [34, 33].

These effects are caused by alterations at the ionic level, which could be summarized by means of the following processes:

i. the reduction of *O*_2_ (*hypoxia*), which causes a slowdown in the process of ATP synthesis. This results in the opening of ATP-sensitive K^+^ channels that induces potassium leakage outside the cell [7, 32];
ii. the accumulation of the potassium ion in the extracellular domain [*K*^+^]_*o*_ (*hyperkalemia*), resulting in the increment of the resting (Nernst) potential *E*_*K*_ [8, 35];
iii. the reduction of intracellular pH (*acidosis*), which decreases the influx of sodium and calcium currents through the cellular membrane [9, 36].

Such ionic and metabolic effects are modeled in this work by starting from the Ten Tusscher-Panfilov model (TTP06) [26], which is one of the most used and exhaustive models for simulating ventricular electrophysiology for 3D numerical experiments. In order to be able to exploit the TTP06 model also in pathological scenarios of AMI, it is fundamental to add to the mathematical ionic model the ATP-sensitive K^+^ current (see point i) above). In recent years, several design of this additional current have been developed (see, e.g., [37, 38, 39, 40, 41, 22, 42, 27]). Specifically, in this study we use the version proposed in [27]:

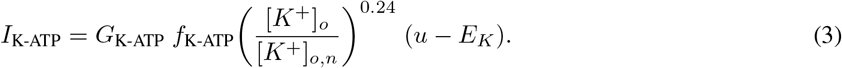

The amplitude of the current depends on the ratio between the actual ([*K*^+^]_*o*_) and the physiological ([*K*^+^]_*o,n*_ = 5.4 *mmol/L*) concentrations. It also depends on the Nernst potential of potassium *E*_*K*_, which varies with [*K*^+^]_*o*_ according to the TTP06 ionic laws, and on the channel conductance *G*_K-ATP_ = 0.05 *mS/μF* [27]. *f*_K-ATP_ is used as scaling factor to vary the conductance in the model, depending on the tissue condition (healthy, transitional or ischemic).

Here, the new current *I*_K-ATP_ is added in two of the equations of the TTP06 model:

- in the potassium dynamics equation, described as the variation in time of the intracellular potassium concentration, calculated as the ratio of the sum of the potassium-dependent currents and *V*_*C*_*F*, representing the product between the cytoplasmic volume and the Faraday constant;
- in the sum of the transmembrane ionic currents.

This modified version of the Ten Tusscher-Panfilov model is useful to represent the metabolic impairments that arise because of the reduced coronary flow [38, 41].

In what follows, we report how the ionic parameters of the ionic TTP06 model are affected by hypoxia, hyperkalemia and acidosis, see [27] for further details. Specifically, the authors proposed to use for the ischemic region characteristic values corresponding to the state occurring ten minutes after the ischemic event, according to experimental cellular curves [33, 43].

The effective choices to model hypoxia, hyperkalemia and acidosis are summarized as follows:

i. the new current conductance is varied through the scaling factor *f*_K-ATP_ as follows: *f*_K-ATP_ = 0 in the *healthy zone* (HZ) so to model the current being silent; *f*_K-ATP_ = 0.1 in the transitional region (*border zone*, BZ); *f*_K-ATP_ = 0.2 in the *ischemic zone* (IZ);
ii. the physiological value of the extracellular concentration of potassium is set [*K*^+^]_*o,n*_ = 5.4 *mmol/L*, and it is increased to [*K*^+^]_*o*_ = 7.2 *mmol/L* in the transitional zone BZ and to [*K*^+^]_*o*_ = 9 *mmol/L* in the ischemic zone IZ;
iii. the reduction of intracellular pH that arises after ten minutes into an ischemic event corresponds to a decrease of the peak *I*_*Na*_ and *I*_*CaL*_ conductances of 25% in IZ. In BZ a reduction of 12% is applied.

In Table 1 we summarize the values of all the physical parameters related to the modified version of the Ten Tusscher-Panfilov model used in this work.

**Table 1:** List of physical parameters used in our study to model hypoxia, hyperkalemia and acidosis, together with their values and references.

|  | Electrophysiology model parameters |  |  | References |
| --- | --- | --- | --- | --- |
| $G_{K-ATP}$ | $0.05 \text{ mS}/\mu\text{F}$ | | | [23] |
|  | Physiological zone | Border zone | Ischemic zone |  |
| $f_{K-ATP}$ | 0 | 0.1 | 0.2 | [23] |
| $[K^+]_o$ | $5.4 \text{ mmol}/L$ | $7.2 \text{ mmol}/L$ | $9 \text{ mmol}/L$ | [43] |
| $G_{Na}$ | $14.838 \text{ mS}/\mu\text{F}$ | $13.057 \text{ mS}/\mu\text{F}$ | $11.1285 \text{ mS}/\mu\text{F}$ | [36] |
| $G_{CaL}$ | $0.0000398 \text{ mS}/\mu\text{F}$ | $0.000035 \text{ mS}/\mu\text{F}$ | $0.00002985 \text{ mS}/\mu\text{F}$ | [36] |

### 2.3 Numerical methods

The electrophysiological model (1) with the modified TTP06 ionic model accounting for (3) is discretized in time with the semi-implicit Backward Differentiation Formula of order 2 (BDF2) [44, 45]. Specifically, the nonlinearities appearing in ***g*** and *I*_*ion*_ are treated by using second order extrapolations of the unknowns *u* and ***w***. Because of the extremely high stiffness of cellular models, due to the significant variables variations over short timescales, a small value for the time-step Δ*t* is required [29]. This also allows us to satisfy the milder absolute stability constraint due to the semi-implicit scheme.

As regards the discretization in space, the transmembrane potential in the left ventricle geometry is modeled by means of the Finite Elements (FE) method. In order to accurately capture the fast propagation of the electrical impulse, quadratic P^2^ FE are employed together with a correspondingly small size of the computational mesh [46]. We used the *Ionic Current Interpolation* (ICI) approach for the ionic current *I*_*ion*_, which builds an interpolation directly in the nodes [47]. This choice is made in order to reduce the computational cost and since we experienced good stability properties.

To provide suitable initial values for the unknowns *u* and ***w***, we perform a pre-processing simulation where the modified TTP06 cellular 0D models (one for each tissue condition, i.e. healthy, transitional, ischemic) are run for about 800 beats until cycle limit values are reached.

### 2.4 Construction of patient-specific configurations

The starting point is the construction of four patient-specific geometries of left ventricles acquired from CT scans of patients suffering from functionally significant coronary artery disease, see [28]. Two acquisitions have been performed for each patient: a *coronary CT angiography* at rest and a myocardial *stress Computed Tomography Perfusion* (stress-CTP) under stress due to adenosine-induced vasodilation. From the first acquisition, three-dimensional geometries of the left ventricle myocardium were reconstructed and turned into a volumetric mesh by using *VMTK*, a collection of libraries and tools for 3D reconstruction, geometric analysis, mesh generation and surface data analysis for image-based modeling [48, 49]; from the second acquisition, the evaluation of myocardial perfusion was achieved through the measurement of myocardial blood flow (MBF) under pharmacologically induced stress conditions. Notice that a perfusion defect in a specific region revealed by stress-CTP indicates that a localized acute ischemia induced by the stress is developing during the procedure.

For each patient, we have at disposal also MBF maps in [*ml/min/*100 *ml*], representing the average value over the heartbeat normalized over 100 *g* of tissue (see Figure 2, top). In order to cluster MBF maps into the three healthy, transitional and ischemic regions, we provide a manipulation of them by means of *Paraview* (open-source, multi-platform data analysis and visualization application based on Visualization Toolkit (VTK) [50]) (see Figure 2, bottom). In accordance with clinical indications, HZ, BZ and IZ in the myocardial tissue are determined as follows:

**Figure 2:**
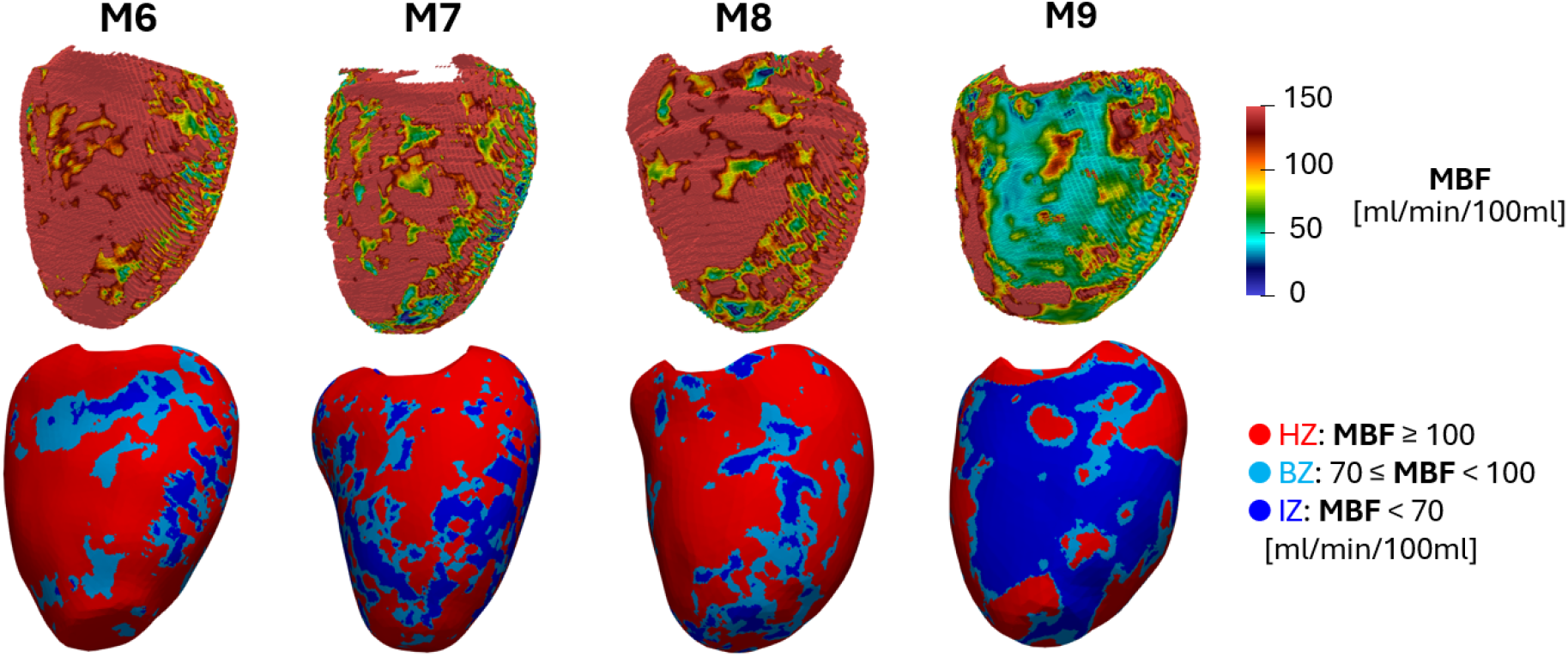
Measurements of Myocardial Blood Flow (MBF) map from stress-CTP (top) and from the clustering manipulation performed with Paraview (bottom). Patients M6, M7, M8, M9 from [28]. Epicardial views. Part I.

- Healthy zone (HZ): MBF values higher than 100 *ml/min/*100 *ml* [51, 52] (red color in the figure);
- Border zone (BZ): in order to account for significant transitional regions [43], we identify the region with MBF values ranging between 70-100 *ml/min/*100 *ml* (light-blue color in the figure);
- Ischemic zone (IZ): MBF values lower than 70 *ml/min/*100 *ml* (blue color in the figure).

According to the values discussed in Section 2.2, we assign to each of these three regions a different value for the variables *f*_K-ATP_, [*K*^+^]_*o*_, *G*_*Na*_, *G*_*CaL*_.

In Table 1 we report the total volumes and the volume percentages of healthy, border and ischemic zones for each patient M6, M7, M8 and M9.

### 2.5 Stimulation procedure

In this work, we consider the following steps to identify the stimulation protocol S⊮:

- The sinus rhythm (SR). To surrogate the propagation of the transmembrane potential from the atrioventricular node to the Purkinje network (not modeled in this work) terminations, three closely spaced in time spherical impulses are triggered at three different coordinates on the left ventricular endocardial surface [53];
- The ectopic event (S1). In order to test the possible induction of arrhythmia, we set a fourth premature impulse (S1) that represents an ectopic event occurring in a vulnerable substrate because of abnormal automaticity or triggered activity [54]. Critical ectopic beats usually take place spontaneously on the epicardial surface along the border zone [55]: in some cases, this abnormal impulse is able to re-excite the tissue at the end of the refractory period of myocardial cells, leading to the formation of reentry mechanisms.

For each patient, we run several simulations applying the S⊮ stimulation protocol. Specifically, we select twelve ectopic sites ***x***_*j*_ that are equispatially distributed along the border zone (see Figure 3) and which are activated at times *t*_*j*_ *> t*^0^, where *t*^0^ is the onset of SR. Among the latter, for each site *j* we look for the ectopic activation time 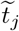 associated to the *Coupling Interval CI*_*j*_, i.e. the smallest distance from *t*^0^ at which the impulse S1 is able to depolarize the tissue without encountering the repolarization front of SR: 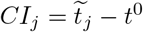.

**Figure 3:**
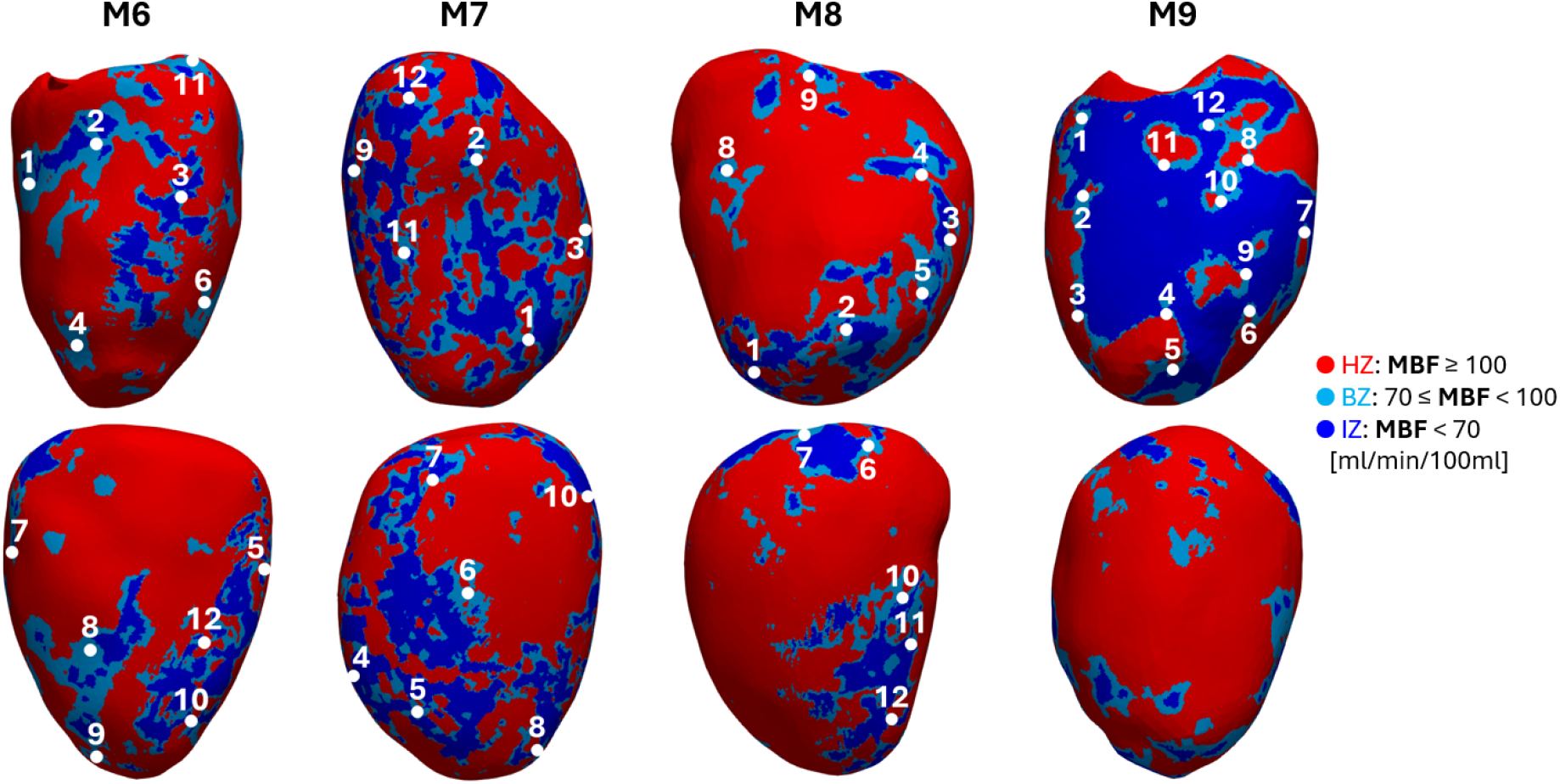
Stimulation sites for the 12 ectopic impulses set for patients M6, M7, M8, M9 from [28]. The two rows depict two different epicardial views of the ventricle. In red the healthy zone (HZ), in light-blue the border zone (BZ), in blue the ischemic zone (IZ). Part I.

To this aim, we propose for each ectopic site ***x***_*j*_ an iterative method where 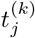 is suitably updated as in Algorithm 1:

#### Algorithm 1

Stimulation procedure to determine CI for the *j* − *th* impulse

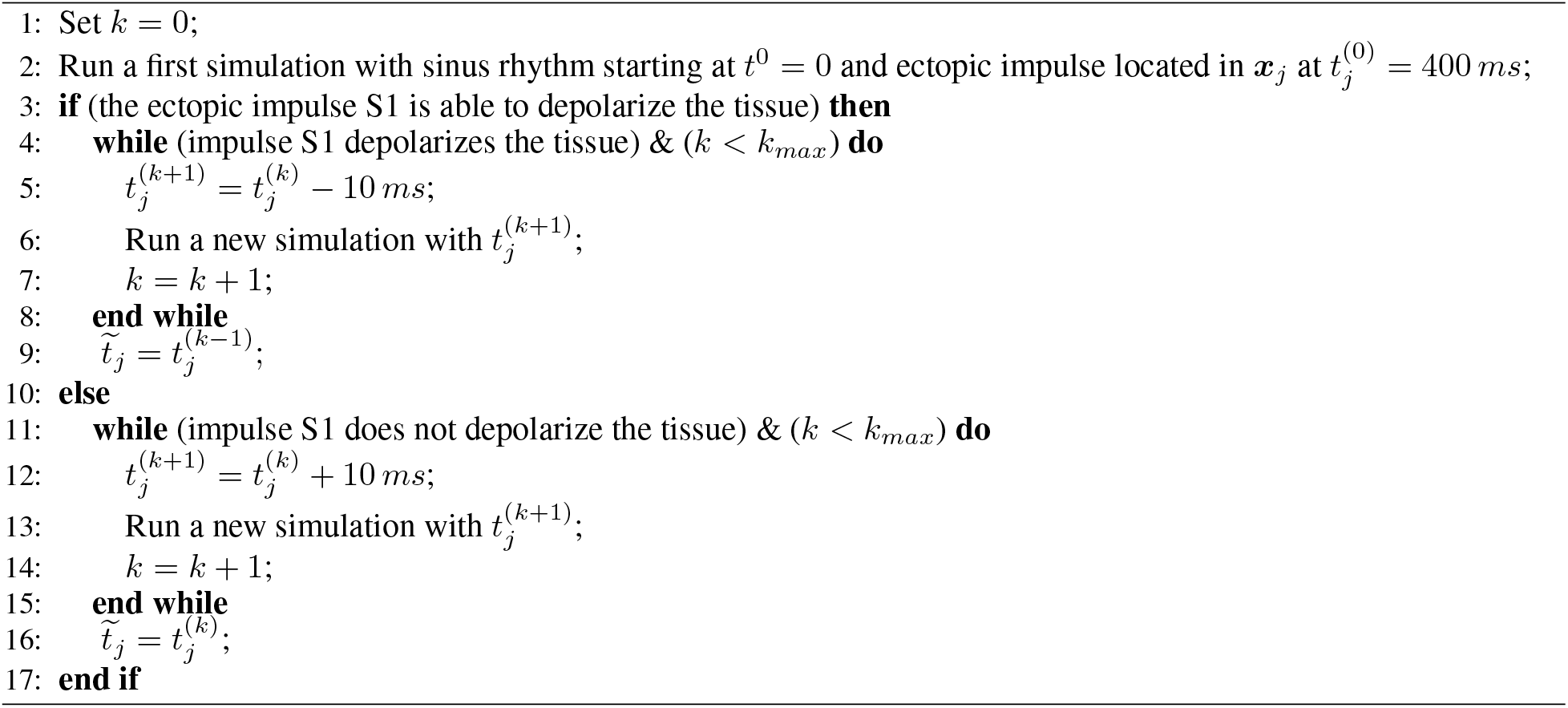

After this procedure, we are able to associate a *CI*_*j*_ to each ectopic site ***x***_*j*_ for each patient. By looking at the outcomes obtained from the final simulation, we count the number of reentry loops that the ectopic impulse is able to generate, so to analyze for each patient the intra-individual arrhythmogenic propensity of the tissue.

### 2.6 Set-up of the numerical experiments

All the numerical experiments are run using `life`^X^ [56], an open-source C++ library for the simulation of the heart functions, developed at MOX, Dipartimento di Matematica, in collaboration with LaBS, Dipartimento di Chimica, Materiali e Ingegneria Chimica “Giulio Natta” (Politecnico di Milano), in particular the repository `life`^X^`-ep` [57]. In this study, a volumetric tetrahedral mesh, with an average edge length *h* = 0.7 *mm* [58] (see Figure 1, middle, where we report as a representative case the computational mesh of patient M9), and a value of the time discretization parameter Δ*t* = 5 · 10^−5^ *s* [59] are used for each patient. Notice that the value of *h* is larger than the usual one used with linear P^1^ FE [29], since in this work we are employing quadratic P^2^ FE, for which the value of *h* could be doubled with equal accuracy [60].

In our simulations we apply spherical impulses for the stimulation conditions, where each impulse has an amplitude of 34.28 *V/s*, a duration of 3 · 10^−3^ *s* and a radius of 6 *mm*, values tuned to generate the propagating action potential. As regards the stimulation procedure used to determine CI, we consider in Algorithm 1 *k*_*max*_ = 40.

As regards the complex fibers-sheets architecture that characterize the myocardium, we employ a Lagrangian-Dirichlet rule-based numerical reconstruction based on the Bayer-Trayanova (BT) model [61]. In order to simulate the traveling impulse in a left ventricle geometry, the monodomain conductivities are calibrated so to obtain a longitudinal conduction velocity equal to 60 *cm/s* (*Σ*_*f*_ = 0.9247 *cm*^2^*/s*, see (2)) and normal and transversal velocities equal to 40 *cm/s* (*σ*_*s*_ = *σ*_*n*_ = 0.3644 *cm*^2^*/s*, see (2)) [62].

Because of the absence of the scarring tissue, no changes of the monodomain conductivities are made to model the pathological scenarios. Indeed, during the early phase of AMI, the reduction of the conduction velocities in the three main directions is only due to the metabolic and ionic heterogeneities that arise in the regions downstream the arterial occlusion: the gap junctions of ventricular muscle are not dissociated yet, so no decrease in coupling conductances is present [11].

## 3 Results Part I - Arrhythmogenesis in presence of patient-specific acute ischemia

In Table 3 we report for all the patients the number of reentry loops *R*_*j*_ that the impulse S1, applied in each ectopic site ***x***_*j*_, is able to generate at time 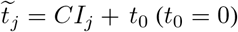, found by means of the proposed Algorithm 1.

As regards patient M6, the ectopic impulses do not generate reentrant activity: after finding *CI*_*j*_ for each ectopic site ***x***_*j*_, we observe that for this patient the impulse, after its onset, is able to propagate and depolarize the myocardial tissue again, but then it decays without forming any reentry loop. We can notice that patient M9 has the highest numbers of reentry loop formation. Indeed, for this patient, S1 is able to generate at least one loop in each ectopic site. Moreover, in points ***x***_8_ and ***x***_9_ the ectopic impulse finds the optimal triggering and electrical remodeling conditions that allow the formation of four reentry loops. By contrast, in patient M7 S1 causes the formation of only one reentry in one ectopic sites over twelve, whereas in patient M8 S1 causes the formation of only one loop in four ectopic sites. Consequently, from our results, patient M9 appear to be the most prone to generate sustained reentrant drivers, thus making it the most arrhythmogenic among the four patients.

In Figure 4 we report the progression of the first three reentries originated in the most arrhythmogenic case M9 for the ectopic impulse located in site ***x***_8_ (identified with a pink circle in row S1) for *CI*_8_ = 420 *ms*. For selected time instants, we show the epicardial view on the left and the endocardial view in the middle, in order to evidence the depolarization maps that arise along the two ventricular surfaces. Moreover, we report a further view of a ventricle’s section on the right, so to highlight the transmural depolarization front. In rows Reentry I, Reentry II and Reentry III, the curved white arrows indicate the formation of reentries at times *t* = 700, 1000, 1350 *ms*, respectively. At times *t* = 550, 900, 1000, 1100, 1350, 1500 *ms* we can notice that the reentrant activity is able to sustain itself because of the propagation in specific regions which occurs only at the endocardium (see the white arrows) and not at the epicardium (see the white circles), since the wavefront is spreading out along the transmural direction. This heterogeneous electrical activation creates the suitable conditions that allow the reentry to form.

**Figure 4:**
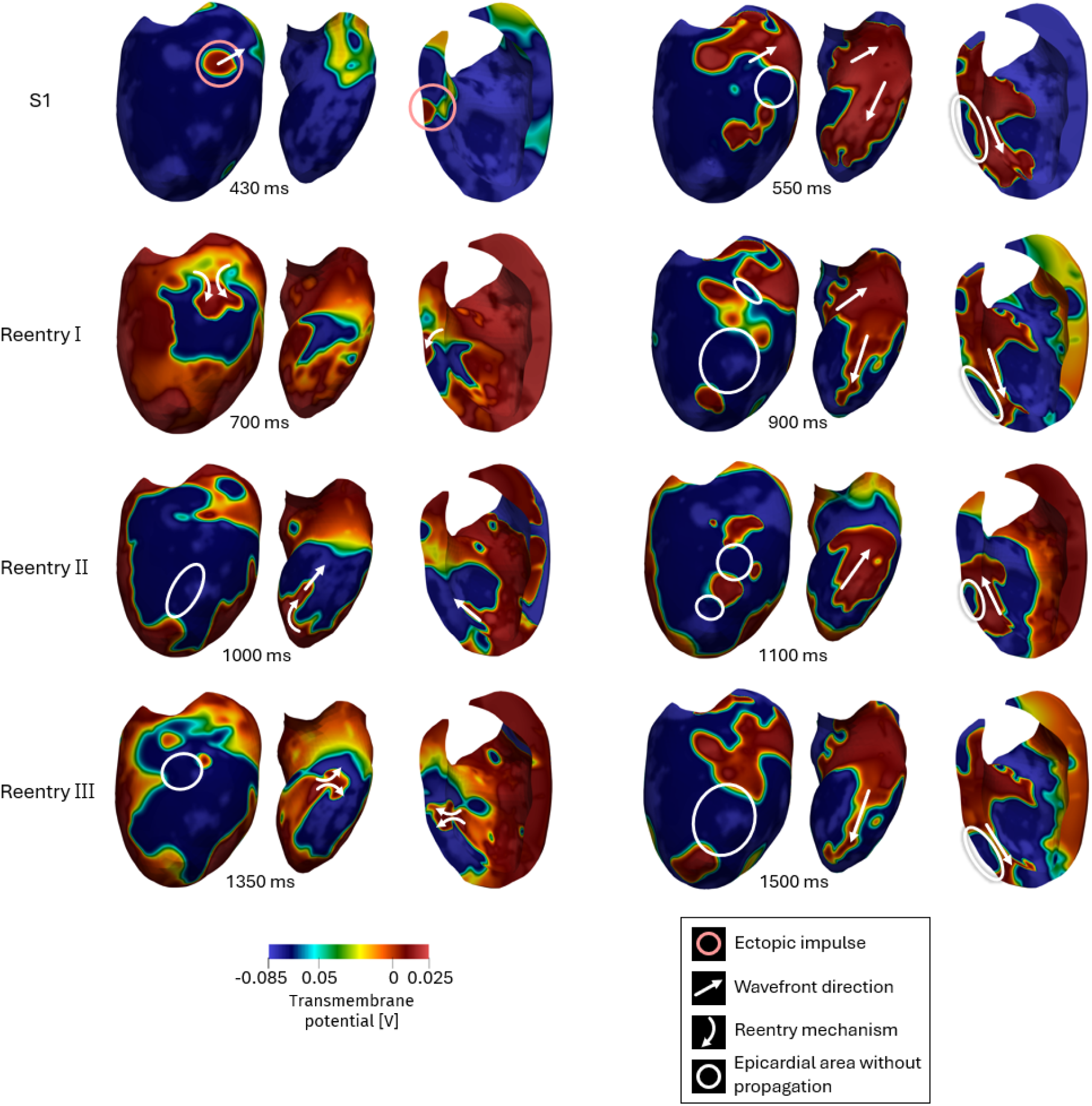
Evolution of reentry loops generated by the ectopic impulse set in site ***x***_8_ at time 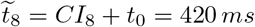 for patient M9. For selected time instants, we report the propagation of the transmembrane potential on the epicardium (left), on the endocardium (middle), and we report a sectional view of the ventricle in which the transmural propagation is visible (right). The first row (S1) represents the propagation just after the ectopic impulse (identified with a pink circle), whereas the other three rows represent the sequence of reentry loops. White straight arrows indicate the direction of the wavefront; white curved arrows indicate part of the reentry circuits; white circles identify the epicardial areas where the wavefront propagation is not allowed. Part I.

In Figure 5, we show for the three least arrhythmogenic cases M6, M7, M8 selected numerical results of maps of the electrical wavefront. Specifically, we report cases M6-***x***_2_ and M7-***x***_3_ (which do not induce any reentry), and M8-***x***_5_ (which induces one reentry loop). In these patients, the ectopic impulses (identified with a pink circle in row S1) are able to depolarize the tissue after being set at time 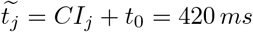, propagating in the directions indicated by white straight arrows. In the three patients, at time *t* = 550 *ms* we can notice a uniform depolarization of the ventricle both at the endocardium and at the epicardium. While in patient M6 and M7 S1 decays without forming any reentry, as we can see at time *t* = 800 *ms*, in patient M8 one reentry (indicated with white curved arrows) is generated at time *t* = 700 *ms* (row Reentry I) at the epicardium, depolarizing the ventricle for the second time.

**Figure 5:**
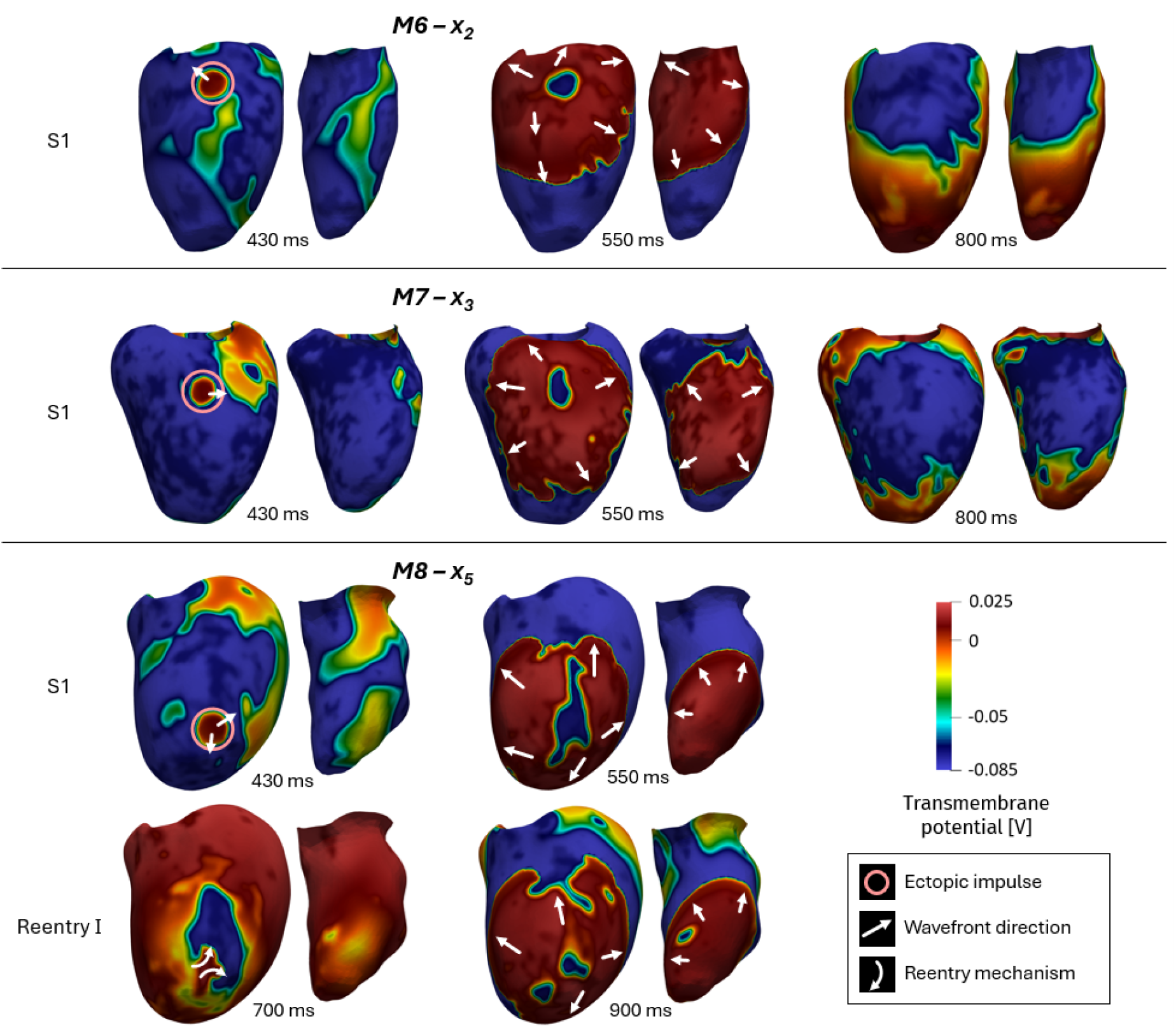
Wavefront progression generated by the ectopic impulses set at time 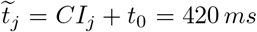 in sites ***x***_2_ for patient M6, ***x***_3_ for patient M7, and ***x***_5_ for patient M8. For selected time instants, we report the propagation of the transmembrane potential on the epicardium (left) and on the endocardium (right). Rows S1 represent the propagation just after the ectopic impulse (identified with a pink circle). Notice the standard repolarization characterizing M6 and M7. On the contrary, row Reentry I in patient M8 represents the formation of a reentry loop. White straight arrows indicate the direction of the wavefront; white curved arrows indicate part of the reentry circuits. Part I.

Referring to Table 2 and considering the outcomes obtained from the simulations just discussed, it is interesting to notice the differences arising between patient M9 and the others. Indeed, although it is the most arrhythmogenic one (see Table 3), this patient turns out not to be the one with larger pathological (border and ischemic) region volumes (see Table 2), which is patient M7.

**Table 2:**
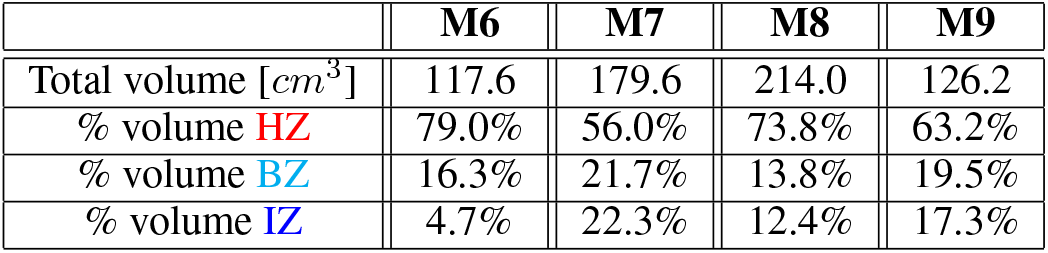
Total myocardial volume and corresponding volume percentages of healthy, border and ischemic zones for each patient M6, M7, M8, M9. Part I.

**Table 3:** Number of reentry loops (*R*_*j*_) that are generated after the onset of the ectopic impulse S1 at times founded with Algorithm 1 for patients M6, M7, M8, M9. Part I.

|  | M6 |  | M7 |  | M8 |  | M9 |  |
| --- | --- | --- | --- | --- | --- | --- | --- | --- |
| j | CI [ms] | R | CI [ms] | R | CI [ms] | R | CI [ms] | R |
| 1 | 370 | 0 | 350 | 0 | 370 | 0 | 380 | 1 |
| 2 | 420 | 0 | 340 | 0 | 420 | 0 | 370 | 1 |
| 3 | 410 | 0 | 420 | 0 | 430 | 0 | 350 | 1 |
| 4 | 390 | 0 | 380 | 0 | 410 | 0 | 370 | 1 |
| 5 | 400 | 0 | 410 | 0 | 420 | 1 | 370 | 1 |
| 6 | 420 | 0 | 400 | 1 | 380 | 0 | 410 | 1 |
| 7 | 390 | 0 | 410 | 0 | 420 | 0 | 390 | 1 |
| 8 | 390 | 0 | 410 | 0 | 390 | 1 | 420 | 4 |
| 9 | 460 | 0 | 410 | 0 | 400 | 1 | 400 | 4 |
| 10 | 370 | 0 | 380 | 0 | 380 | 0 | 410 | 1 |
| 11 | 420 | 0 | 410 | 0 | 380 | 1 | 390 | 1 |
| 12 | 390 | 0 | 400 | 0 | 370 | 0 | 420 | 1 |

## 4 Methods Part II - Sensitivity analysis of acute ischemic regions

In order to evaluate how the extension and the volume of an AMI region can influence the generation of reentrant mechanisms in pathological left ventricles, we create two virtual scenarios of AMI for patient M9, the one that showed the highest arrhythmic propensity. The same mathematical and numerical methods explained in Sections 2.1, 2.2, 2.3 are used to run the simulations in `life`^X^. Starting from the patient-specific geometry of patient M9 (Section 2.4), together with the clinical range (reported in the bullet points in Section 2.4 and, in what follows, referred to as *SEVERE*, SE), we consider two further ranges of MBF to describe two smaller ischemic and border zones (see Figure 6):

**Figure 6:**
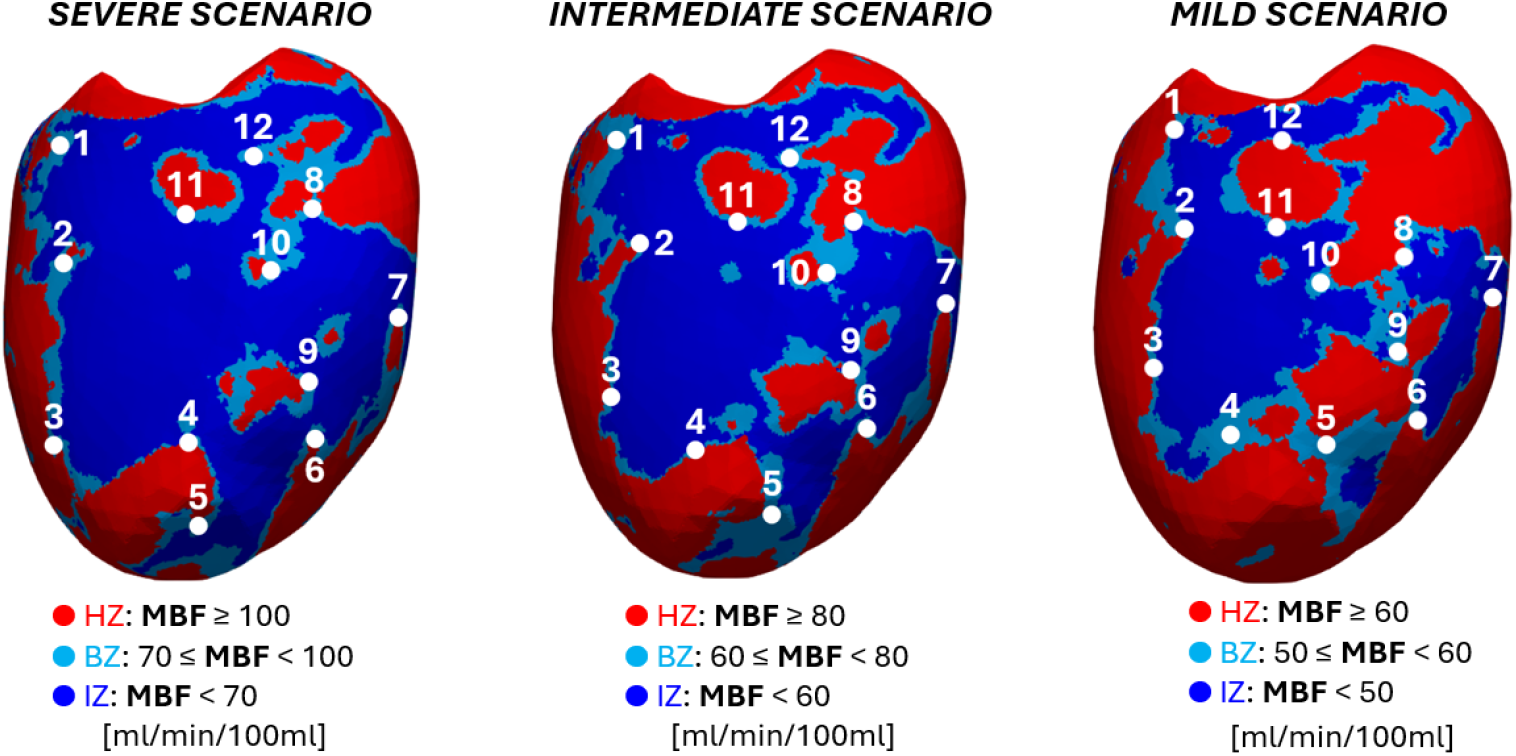
Stimulation sites for the 12 ectopic impulses set for patient M9 (left, SEVERE scenario (SE)) and for the two virtual scenarios (center, INTERMEDIATE scenario (IN); right, MILD scenario (MI)). In red the healthy zone (HZ), in light-blue the border zone (BZ), in blue the ischemic zone (IZ). Part II.

- Scenario *INTERMEDIATE* (IN):
  - HZ: MBF values higher than 80 *ml/min/*100 *ml* (red color in Figure 6);
  - BZ: MBF values ranging between 60-80 *ml/min/*100 *ml* (light-blue color);
  - IZ: MBF values lower than 60 *ml/min/*100 *ml* (blue color).
- Scenario *MILD* (MI):
  - HZ: MBF values higher than 60 *ml/min/*100 *ml* (red color);
  - BZ: MBF values ranging between 50-60 *ml/min/*100 *ml* (light-blue color);
  - IZ: MBF values lower than 50 *ml/min/*100 *ml* (blue color).

These scenarios can further highlight how the distribution of the pathological regions influences the formation of reentries. In Table 4 we report the volume percentages of healthy, border and ischemic zones for the three scenarios considered for M9.

**Table 4:** Volume percentages of healthy, border and ischemic zones for SEVERE (SE), INTERMEDIATE (IN), and MILD (MI) scenarios of M9. Part II.

|  | SE | IN | MI |
| --- | --- | --- | --- |
| % volume HZ | 63.2% | 76.4% | 87.5% |
| % volume BZ | 19.5% | 11.1% | 4.5% |
| % volume IZ | 17.3% | 12.5% | 8.0% |

In order to simulate a possible case of spontaneous ectopic impulse, we use again the protocol described in Section 2.5 and the same set-up presented in Section 2.6. The twelve new ectopic sites ***x***_*j*_ in the two virtual scenarios are also distributed along the border region as close as possible to the points chosen for the SE case to eliminate from the analysis the counfounding factor given by the influence of the locations of the ectopic impulse on arrhythmogenesis.

## 5 Results Part II - Sensitivity analysis of acute ischemic regions

In Table 5 we report for SEVERE (SE), INTERMEDIATE (IN) and MILD (MI) scenarios of patient M9 the number of reentry loops *R*_*j*_ that the impulse S1, applied in each ectopic site ***x***_*j*_, is able to generate at time 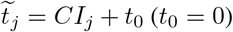, found by means of the proposed Algorithm 1.

**Table 5:** Number of reentry loops (*R*_*j*_) that are generated after the onset of the ectopic impulse S1 at times founded with Algorithm 1 for SEVERE (SE), INTERMEDIATE (IN) and MILD (MI) scenarios. Part II.

|  | SE |  | IN |  | MI |  |
| --- | --- | --- | --- | --- | --- | --- |
| j | CI [ms] | R | CI [ms] | R | CI [ms] | R |
| 1 | 380 | 1 | 380 | 1 | 390 | 1 |
| 2 | 370 | 1 | 360 | 1 | 370 | 1 |
| 3 | 350 | 1 | 350 | 0 | 340 | 0 |
| 4 | 370 | 1 | 380 | 1 | 370 | 1 |
| 5 | 370 | 1 | 380 | 1 | 390 | 0 |
| 6 | 410 | 1 | 380 | 1 | 410 | 1 |
| 7 | 390 | 1 | 390 | 0 | 400 | 0 |
| 8 | 420 | 4 | 420 | 2 | 410 | 0 |
| 9 | 400 | 4 | 400 | 1 | 400 | 1 |
| 10 | 410 | 1 | 400 | 1 | 400 | 0 |
| 11 | 390 | 1 | 400 | 0 | 400 | 1 |
| 12 | 420 | 1 | 420 | 0 | 410 | 0 |

We can notice that the SE scenario is able to generate at least one reentry loop for each ectopic impulse. Instead, for IN and MI scenarios, one reentry occurs in seven and six ectopic sites, respectively, whereas two loops only in one case for IN. Consequently, as expected, our results highlight that the SE scenario is more arrhythmogenic than IN and MI.

To emphasize the different propagation patterns occurred in the three scenarios, we focus our analysis on the case of the ectopic impulse activated in site ***x***_8_. By looking at Figure 4 for SE and at Figure 7 for IN and MI, where with the pink circle we indicate the location of the ectopic impulse, it is possible to see that, in the first instants after the onset of the impulse (*t* ≤ 550 *ms*, row S1), the depolarization fronts of all the three scenarios are very similar and propagate along the same direction (identified by white straight arrows).

**Figure 7:**
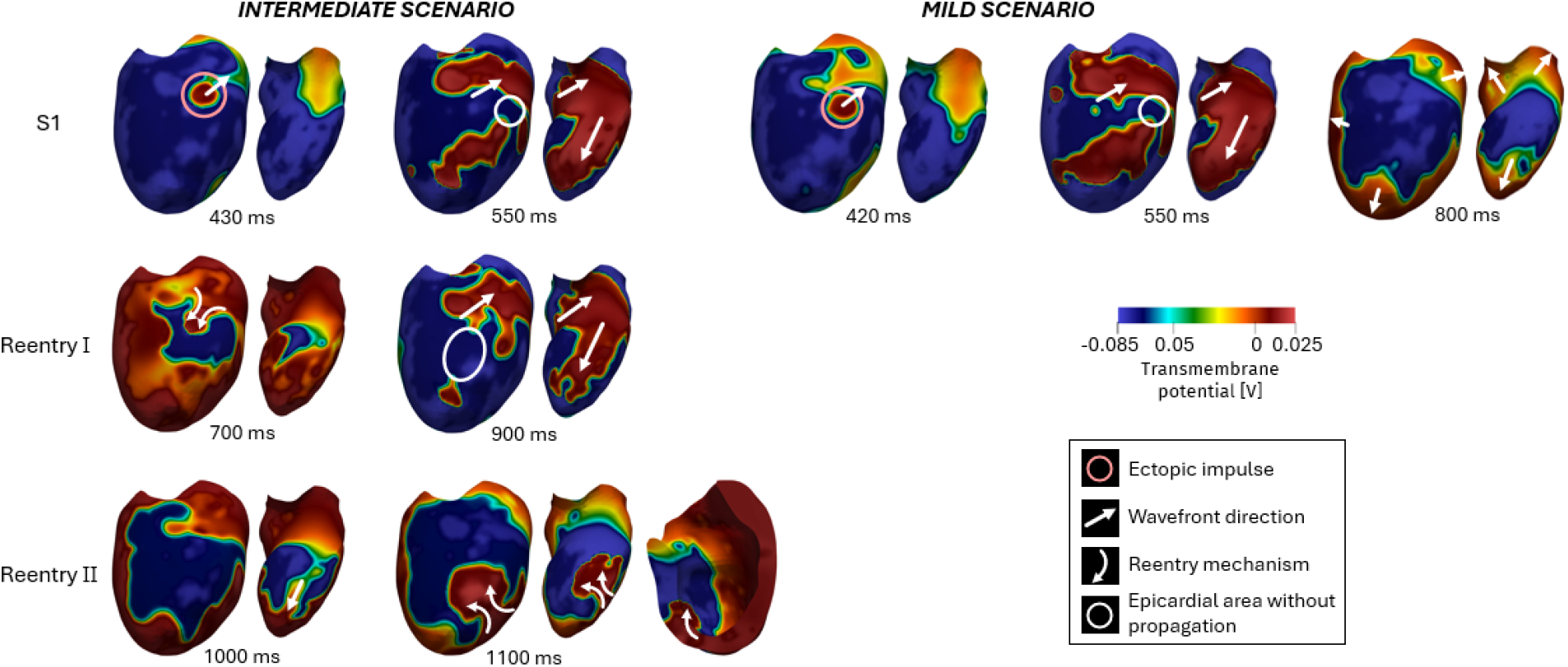
Evolution of reentry loops generated by the ectopic impulse set in sites ***x***_8_ at time 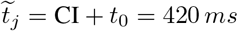 for INTERMEDIATE (IN) scenario and at time 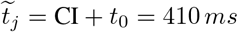 for MILD (MI) scenario of patient M9. For selected time instants, we report the propagation of the transmembrane potential on the epicardium (left) and on the endocardium (right); moreover, for IN we report a sectional view of the ventricle in which the transmural propagation is visible at time *t* = 1100 *ms*. The first row (S1) represents the propagation just after the ectopic impulse (identified with a pink circle), whereas the other two rows represent the sequence of reentry loops. White straight arrows indicate the direction of the wavefront; white curved arrows indicate part of the reentry circuits; white circles identify the epicardial areas in which the wavefront propagation is not allowed. Part II.

Going further in the analysis of the simulations’ results, for 700 *ms* ≤ *t* ≤ 900 *ms* we observe different behaviors of the three scenarios. Specifically, for MI the ectopic impulse decays without generating any reentry mechanisms; instead, for IN and SE we observe a very similar propagating wavefront (see row Reentry I): in a similar way to that explained in Section 3, also for IN a first reentry is generated (see *t* = 700 *ms*) and endocardial pathways allow the sustainment of the propagation of the wavefront (see *t* = 900 *ms*).

Instead, by looking at what happens for 1000 *ms* ≤ *t* ≤ 1100 *ms*, we can observe that both IN and SE generate a second reentry, however with very different mechanisms and patterns. Specifically, unlike the SE case, for IN such reentry is present both at endocardium and epicardium (see *t* = 1100 *ms*) and thus throughout the entire thickness of the myocardial region (see the sectional view reported on the right at time *t* = 1100 *ms*). This does not create the heterogeneous electrical condition which would allow a further (third) reentrant loop. Indeed, after the second reentry loop, the tissue repolarizes.

## 6 Discussion

### 6.1 On the novelties of the work

In this work, we report (at the best of our knowledge) the first computational study which includes patient-specific information on AMI regions, to assess the influence of Acute Myocardial Ischemia (AMI) on arrhythmogenesis. The use of patient-specific information on AMI distributions is fundamental to accurately assess the risk of arrhythmia formation in the specific patient. Indeed, the use of scar distribution as a surrogate of AMI zones [25] may not be very informative, since changes occurring at the ionic level during the early acute ischemic phase do not necessarily evolve into an infarct.

Accordingly, the main novelty of this work is to employ Myocardial Blood Flow (MBF) maps, coming from stress-CTP exams, in numerical experiments of four ischemic cases. Indeed, during this type of acquisition, a sudden and reversible acute ischemia is pharmacologically induced.

### 6.2 Propensity of arrhythmogenesis: inter- and intra-patients analyses

We first discuss the results obtained from the comparison among patients performed in Section 3. Our method was based on selecting multiple ectopic sites on the border regions covering a wide portion of the epicardium, unlike previous studies where only a single ectopic stimulus was often considered [22, 25]. This allows us to explore throughout the entire ventricular myocardium the tendency of the patients to be arrhythmogenic.

Moreover, the inter-individual analysis allows us to examine the impact of the volume and of the distribution of border zone (BZ) and ischemic zone (IZ) on arrhythmogenesis. Indeed, MBF maps acquired from four patients with different severity and types of stenosis show heterogeneous pathological areas, characterizing the left ventricles with very dissimilar ischemic regions. It is noticeable in Figure 3 that patient M9 presents a more confined ischemic area with respect to the other three, which show a scattered ischemic configuration. Moreover, by performing an analysis on the volume percentages of the regions, as reported in Table 2, it is interesting to notice that the patient with bigger BZ and IZ is M7, followed by M9, M8 and finally M6. Consequently, from the outcomes presented in Section 3, where M9 results to be the most arrhythmogenic patient, we can suggest that the property of the ischemic region that has a major influence on arrhythmogenesis is not only its volume, but also its heterogeneous distribution throughout the myocardium.

A crucial aspect of patient M9 is that its arrhythmic propensity is accentuated by the presence of transmural and endocardial healthy pathways that allow the propagation of the impulse, as we can see in Figure 4. This is due to an heterogeneous distribution of ischemia along the myocardial thickness. Owing to this epi-endocardial heterogeneity, the depolarization front is able to pass under epicardial regions that are still in their refractory state, thus reaching areas that can be depolarized and thus enabling the sustainment of the reentrant activity. This transmural heterogeneity could be due to the presence of a wash-out zone in the endocardium, which results from the interaction between the endocardial surface and the ventricular cavity. Indeed, thanks to this connection, oxygen and nutrients are able to diffuse from the cavity to the tissue, ensuring higher perfusion levels in the endocardium with respect to the epicardium. This condition is known to be arrhythmogenic during the first minutes of ischemia [63].

The second part of this study was devoted to a parametric study to assess the influence of the dimension of AMI regions on arrhythmogenesis, whose results are reported in Section 5. To this aim, we studied the arrhythmic tendency of two virtual scenarios (INTERMEDIATE - IN and MILD - MI) of patient M9 which are healthier than the original one (identified here as SEVERE - SE). We select this patient because it showed the highest number of reentry loops in most ectopic sites.

What we observed is that, with respect to SE case, IN and MI scenarios are less prone to generate arrhythmias, as highlighted by Table 5. Specifically, the pro-arrhythmic propensity able to sustain the reentries decreases when BZ and IZ volumes (Table 4) decrease: in such a condition the ectopic impulse propagates with a higher conduction velocity, fastly depolarizing the tissue and hence encountering earlier the refractory state of the pathological regions, which blocks the wavefront.

It is interesting to correlate the values of volume percentages, reported in Table 2 and Table 4, with the corresponding number of reentries, reported in Table 3 and Table 5. We can notice that the regions’ volumes of IN scenario are comparable with those of patient M8, and that MI scenario has the most extended healthy zone among all scenarios/patients. However, patient M8 is less arrhythmogenic than IN: in M8 the impulse is able to generate one reentry only in four ectopic sites over twelve; by contrast, in IN the impulse is able to generate at least one reentry loop in eight ectopic sites over twelve. This observation is also confirmed by comparing M6 and MI: even though M6 has a less extended healthy zone with respect to MI, is is less arrhythmogenic. This outcome confirms that the volume percentage of AMI regions seems to have less influence on arrhythmogenesis than their distribution inside the ventricle.

In conclusion, this work represents a first step toward mechanistic understanding of the patient-specific impact of the distribution of poorly perfused regions within different left ventricles affected by acute ischemia on the onset of arrhythmias. However, it is important to point out that this study does not aim to create a digital twin of a case of acute myocardial ischemia: the early phases of this pathology are hardly measurable in clinic because of the impossibility to obtain immediate data, unless the patient is already in the hospital. For this reason, we introduced an innovative way to consider patient-specific AMI regions by employing ventricles of patients in which an acute ischemic event is pharmacologically induced during stress-CTP.

### 6.3 Limitations and future developments

This work presents some limitations.

- First, the model does not include the His-Purkinje System (HPS). This is important to obtain an accurate conduction of the impulse with the correct sites and times of activation inside the ventricle [64]. However, in view of our comparisons, we believe that our results could provide useful preliminary indications in patient-specific studies of arrhythmogenesis induced by AMI. Future studies on the inclusion of HPS will be however mandatory to follow up on what has been done so far.
- The transitional values of the electrophysiological parameters set in the border zone are kept constant and equal to an intermediate value between the values in the healthy and the ischemic zones. This does not reflect what is shown from experiments [43], that is a linear variation from healthy to ischemic zones.
- The ventricular domain is composed by uniform myocardial properties and the same ionic model is considered for all the cells. Thus, there is no transmural and apex-to-base heterogeneity in the action potential duration [65, 20]. Further analyses should be conducted on such points in order to investigate their influence on arrhythmogenesis.
- Finally, the use of more complex stimulation protocols should be considered. Indeed, in real cases of ischemic ventricles, the activation of a single ectopic impulse hardly sustains an arrhythmic event: several ectopic impulses can originate at high frequencies one after the other in a single activation site [55].

## Data Availability

All data produced in the present study are available upon reasonable request to the authors.

## 7 Acknoledgments*

AC, SP, CV are members of the INdAM group GNCS “Gruppo Nazionale per il Calcolo Scientifico” (National Group for Scientific Computing). CV is partially supported by the Italian Ministry of University and Research (MIUR) within the PRIN (Research projects of relevant national interest) MIUR PRIN22-PNRR n. P20223KSS2 “Machine learning for fluid-structure interaction in cardiovascular problems: efficient solutions, model reduction, inverse problems”. CV and SP are partially supported by the Italian Ministry of Health within the PNC PROGETTO HUB LIFE SCIENCE - DIAGNOSTICA AVANZATA (HLS-DA) “INNOVA”, PNC-E3-2022-23683266–CUP: D43C22004930001, within the “Piano Nazionale Complementare Ecosistema Innovativo della Salute” - Codice univoco investimento: PNC-E3-2022-23683266. AC is funded by INNOVA. SP acknowledges the support of the MUR, Italian Ministry of University and Research, grant Dipartimento di Eccellenza 2023–2027 and of the INdAM GNCS project CUP E53C23001670001.

